# Systematic benefit-risk assessment for the use of chloroquine or hydroxychloroquine as a treatment for COVID-19: Establishing a dynamic framework for rapid decision-making

**DOI:** 10.1101/2020.05.07.20093989

**Authors:** Vicki Osborne, Miranda Davies, Sandeep Dhanda, Debabrata Roy, Samantha Lane, Alison Evans, Saad Shakir

## Abstract

**Objectives:** Given the current pandemic, there is an urgent need to identify effective, safe treatments for COVID-19 (coronavirus disease). A systematic benefit-risk assessment was designed and conducted to strengthen the ongoing monitoring of the benefit-risk balance for chloroquine (CQ) and hydroxychloroquine (HCQ) in COVID-19 treatment.

**Methods:** The overall benefit-risk of the use of chloroquine or hydroxychloroquine as a treatment for COVID-19 compared to standard of care, placebo or other treatments was assessed using the Benefit-Risk Action Team (BRAT) framework. We searched PubMed and Google Scholar to identify literature reporting clinical outcomes in patients taking chloroquine or hydroxychloroquine for COVID-19. A value tree was constructed and key benefits and risks were ranked by two clinicians in order of considered importance.

**Results:** Several potential key benefits and risks were identified for use of hydroxychloroquine or chloroquine in COVID-19 treatment. Currently available results did not show an improvement in mortality risk; Cox proportional hazard ratio (HR) for death between patients who received HCQ alone vs. neither hydroxychloroquine or azithromycin was 1.08 (95% CI 0.63-1.85). A further study compared the incidence of intubation or death (composite outcome) in a time to event analysis between patients who received HCQ vs. those patients who did not (adjusted Cox proportional HR 1.00 (95% CI 0.76-1.32)). Risk of cardiac arrest, abnormal electrocardiogram (ECG) and QT prolongation was greater among patients taking HCQ (with or without azithromycin) compared to standard of care in the same study.

**Conclusions:** Overall, based on the available data there does not appear to be a favourable benefit-risk profile for chloroquine or hydroxychloroquine compared to standard of care in treatment of severe COVID-19. As further data from clinical trials and real world use on these benefits and risks becomes available, this can be incorporated into the framework for an ongoing benefit-risk assessment.

## 1 Introduction

Coronaviruses have circulated among humans and animals for many years, of which several strains are highly transmissible and pathogenic in humans [1]. Severe Acute Respiratory Syndrome (SARS) emerged in 2002 and 2003, while Middle East Respiratory Syndrome coronavirus (MERS-CoV) emerged 10 years later [1]. In December 2019, a novel coronavirus emerged in Wuhan, China [2], subsequently called Severe Acute Respiratory Syndrome coronavirus 2 (SARS-CoV-2) [3]. SARS-CoV-2 causes coronavirus disease (COVID-19) [3] and the outbreak was declared a pandemic by the World Health Organisation (WHO) in March 2020 [4].

Coronaviruses predominantly cause respiratory tract infections in humans [1]. Specifically, the main symptoms of COVID-19 have been reported as fever, cough and shortness of breath [5], with a less abrupt onset of symptoms compared to SARS [6,7]. Data are still emerging regarding the epidemiology of COVID-19, though initial reports estimate a transmission rate (basic reproduction number, R_0_) of 2.2 [8] and a case fatality rate that increases among older adults [9]. Given the current pandemic, there is an urgent need to identify effective, safe treatments for COVID-19. Two such proposed treatments are chloroquine and hydroxychloroquine, which are well-established medications predominantly used to treat malaria, lupus and rheumatoid arthritis. In vitro studies have shown that chloroquine and hydroxychloroquine are effective at inhibiting SARS-Cov-2 infection, with the latter appearing to have more potent antiviral activity [10,11]. Thus, repurposing of these drugs as antiviral therapies for COVID-19 is of global interest, however clinical data are limited and inconclusive. Currently, there are multiple ongoing clinical trials for use of these treatments in COVID-19, while the US Food and Drug Administration (FDA) has issued an Emergency Use Authorization (EUA) for oral formulations of chloroquine phosphate and hydroxychloroquine sulfate for patients hospitalised with COVID-19 [12]. To date, whilst there have been many publications which have described the main effectiveness and safety concerns with these treatments, there has not been a systematic benefit-risk assessment on the use of chloroquine or hydroxychloroquine for COVID-19 treatment using a structured descriptive framework.

A systematic benefit-risk assessment strengthens the ongoing monitoring of the benefit-risk balance for chloroquine or hydroxychloroquine in COVID-19 treatment. For this assessment, the Benefit-Risk Action Team (BRAT) framework is highly applicable as it allows identification of the key benefits and risks of a product in a defined disease context within a structured descriptive framework; further quantitative assessments can then be applied and conducted according to the availability of relevant data at that time [13]. This dynamic approach to benefit-risk assessment of potential treatments for COVID-19 has been previously applied to the anti-viral agent remdesivir [14]. The BRAT framework is also specifically designed to assist communication with regulatory authorities [15]. The decision-making process is transparent due to the framework design, while any assumptions can be explored further by sensitivity analysis through a quantitative component [16].

Given the public health urgency with the COVID-19 pandemic, this benefit-risk assessment has been conducted based on publicly available information to date (data-lock May 27^th^ 2020). It is however acknowledged that there is limited data available from ongoing clinical trials at this timepoint. To inform the debate expeditiously the benefit-risk assessment has been designed to be implemented regardless of the quantity of data available. The intention is that the framework will subsequently be readily available to repeat the assessment as further data arise, e.g. results from new clinical trials, allowing for rapid decision-making.

## 2 Objectives

To examine the benefit-risk profile of chloroquine or hydroxychloroquine in COVID-19 patients compared to standard of care, placebo or other treatments.

## 3 Methods

### 3.1 Benefit-Risk Framework

The overall benefit-risk of the use of chloroquine or hydroxychloroquine as a treatment for COVID-19 compared to standard of care, placebo or other treatments was assessed using the BRAT framework. BRAT uses a six step iterative process to support the decision and communication of a Benefit-Risk Assessment: define decision context, identify outcomes, identify data sources, customise framework, assess outcome importance, and display and interpret key Benefit-Risk metrics [15,16]. Three settings of interest were identified for use of these treatments in COVID-19; treatment for severe disease, treatment of mild disease in the community, and prevention in health care professionals exposed to the virus. For the purposes of this benefit risk assessment, we have focused on the use of chloroquine or hydroxychloroquine for the treatment of severe COVID-19 disease. We defined severe disease as those patients who were hospitalised with COVID-19.

#### 3.1.1 Population of interest

The population of interest were patients with severe COVID-19, while the exposure of interest was hydroxychloroquine or chloroquine. The comparators of interest were standard of care, placebo or other treatments for COVID-19.

#### 3.1.2 Outcomes of interest

Initially, all potential benefits and risks related to hydroxychloroquine or chloroquine, regardless of importance, were identified. All available data sources were used in this process, including case series and case reports from published literature. From these the key benefits and risks associated with hydroxychloroquine and chloroquine use were selected by two clinicians. Key benefits and risks were those which were considered to drive the benefit-risk balance of the drug. A value tree was constructed using these key benefits and risks, all of which were ranked in order of considered importance.

#### 3.1.3 Data sources and customisation of the framework

We searched PubMed and Google Scholar to identify suitable data for inclusion. In both databases, we searched for papers on:

> (((((((((chloroquine* AND SARS*)) OR (chloroquine* and covid*)) OR (chloroquine* AND coronavirus))))) OR (((chloroquine* AND 2019-NCov))))
>
> (((((((((hydroxychloroquine* AND SARS*)) OR (hydroxychloroquine* and covid*)) OR (hydroxychloroquine* AND coronavirus))))) OR (((hydroxychloroquine* AND 2019-NCov))))

Papers were included in the data extraction tables if they reported quantitative data on effectiveness and/or safety of chloroquine or hydroxychloroquine and a comparator in patients with severe COVID-19. Case reports in the published literature were excluded from the data extraction tables. Results were restricted to English language only (abstracts in English language were acceptable where sufficient data provided) and peer-reviewed publications since 2019 to 29^th^ April 2020. An updated search in PubMed only was also conducted to May 27^th^ 2020. Data were extracted for each benefit and risk, for hydroxychloroquine, chloroquine and the comparator (standard of care, placebo or other treatments), where available. EudraVigilance (up to 4^th^ April 2020) and FAERS spontaneous reporting data (up to 31^st^ December 2019) for hydroxycholoroquine and chloroquine were also examined, but were not included in the data extraction tables due to lack of a comparator.

### 3.2 Outcome assessment

A summary benefit-risk table was created to allow visualisation of the magnitude of each benefit and risk. Risk differences and corresponding 95% confidence intervals (CI) were calculated for each outcome where both numerator (number of events) and denominator (number of patients at risk) were available for both the treatment group (hydroxychloroquine or chloroquine) and comparator group. Spontaneous reports are not included in the benefit-risk table and are presented in the text only.

#### 3.2.1 Quantitative assessment

Due to lack of data, a fully quantitative assessment was not undertaken. Risk per 1000 patients, risk differences and 95% confidence intervals (CI) were calculated. However, the outcomes identified in the value tree were ranked so that swing weighting can be applied in future assessments. The weighted net clinical benefit (wNCB) can subsequently be calculated using these weights [16–18]. We would propose using the Sutton et al method, where benefits have a positive contribution to the wNCB and risks have a negative contribution [18]; the overall wNCB would be considered positive (benefit outweighs the risk) where wNCB >0. A sensitivity analysis can also be used to examine the robustness of the assigned weights and whether significant changes would alter the benefit-risk profile [13]. The wNCB was not calculated due to limited data for the current assessment; no data were identified for several key benefits and risks.

## 4 Results

The value tree reflecting the key benefits and risks related to hydroxychloroquine or chloroquine treatment in COVID-19 is displayed in Fig 1. Data for these outcomes are presented in the data extraction table and key benefit-risk summary table (Tables 1 and 2, respectively). From literature searching we identified 238 papers from PubMed and 946 results from Google Scholar for chloroquine. We also identified 403 papers from PubMed and 648 results from Google Scholar for hydroxychloroquine. After initial review and removal of duplicates, 18 papers were reviewed further to determine if they met all inclusion criteria; eight papers were included in the final benefit-risk assessment.

**Fig 1.**
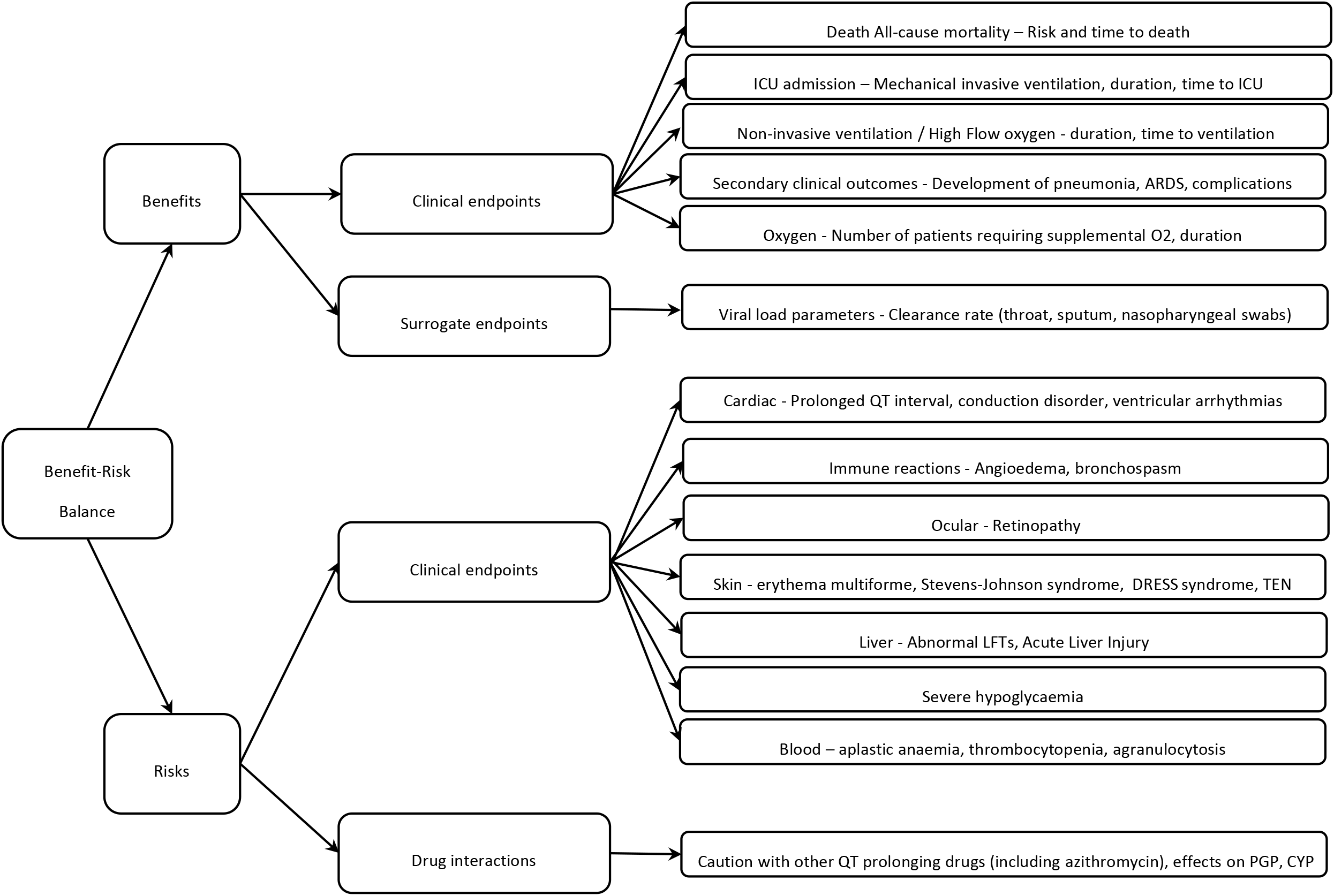
Value tree of key benefits and risks identified for chloroquine and hydroxychloroquine in context of treatment of severe COVID 19 disease

**Table 1.**
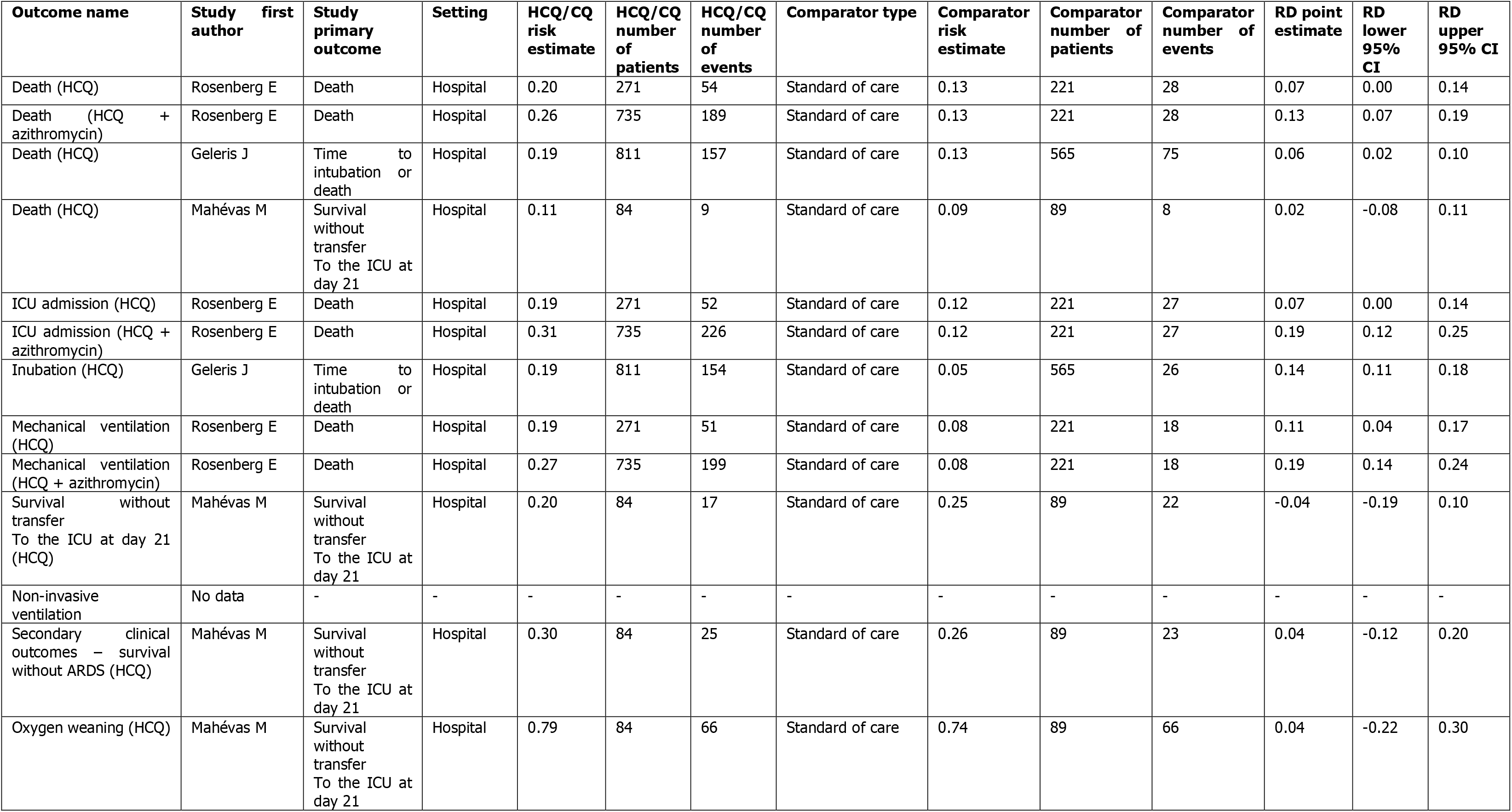
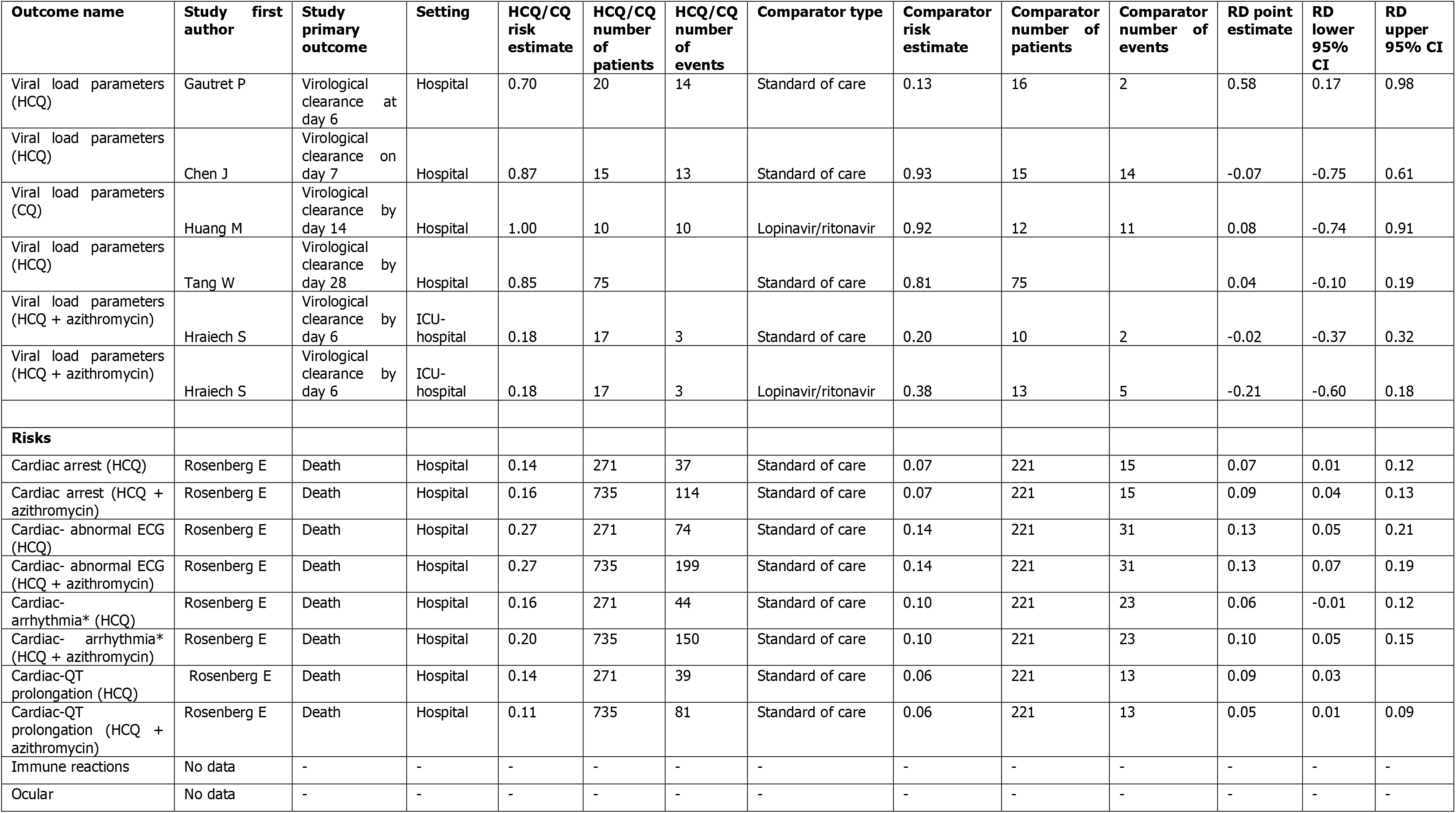
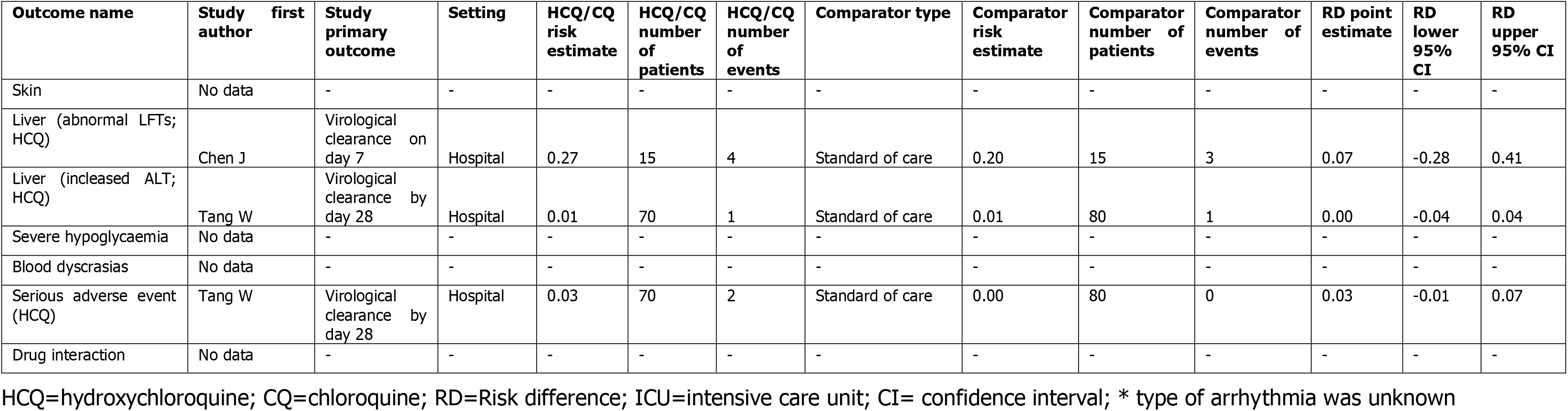
Data for key benefits and risks identified for hydroxychloroquine and chloroquine from peer-reviewed, published literature

**Table 2.** Benefit-Risk summary table for key benefits and risks identified for hydroxychloroquine and chloroquine alone compared to standard of care

| Outcome name | HCQ/CQ risk/1000 patients | Comparator risk/1000 patients | RD (95% CI)/1000 patients | Hazard ratio (95% CI) |
| --- | --- | --- | --- | --- |
| **Benefits** |  |  |  |  |
| Death (Rosenberg et al; HCQ) | 199 | 127 | 73 (2, 143) | 1.08 (0.63, 1.85) |
| Death (Geleris et al; HCQ) | 194 | 133 | 61 (18, 104) |  |
| Death (Mahévas et al; HCQ) | 107 | 90 | 17 (-76, 111) | 1.2 (0.5, 3.0) |
| ICU admission (Rosenberg et al; HCQ) | 191 | 122 | 70 (0, 139) |  |
| Mechanical ventilation (Rosenberg et al; HCQ) | 188 | 81 | 107 (43, 171) |  |
| Intubation (Geleris et al; HCQ) | 190 | 46 | 144 (109, 179) |  |
| Survival without transfer To the ICU at day 21 (Mahévas et al; HCQ) | 202 | 247 | -45 (-186, 96) | 0.8 (0.4, 1.5) |
| Survival without ARDS (Mahévas et al; HCQ) | 298 | 258 | 40 (-118, 197) | 1.2 (0.7, 2.2) |
| Oxygen weaning (Mahévas et al; HCQ) | 786 | 742 | 44 (-217, 305) | 1.1 (0.9, 1.3)^ |
| Viral load parameter- virological clearance at day 6 (Gautret et al; HCQ) | 700 | 125 | 575 (169, 981) |  |
| Viral load parameter- virological clearance at day 7 (Chen et al; HCQ) | 867 | 933 | -67 (-746, 612) |  |
| Viral load parameter- virological clearance at day 28 (Tang et al; HCQ) | 854 | 813 | 41 (-103, 185) |  |
| **Risks** |  |  |  |  |
| Cardiac arrest (Rosenberg et al; HCQ) | 137 | 68 | 69 (13, 124) |  |
| Cardiac- abnormal ECG (Rosenberg et al; HCQ) | 273 | 140 | 133 (53, 212) |  |
| Cardiac- arrhythmia (Rosenberg et al; HCQ) | 162 | 104 | 58 (-6, 122) |  |
| Cardiac- QT prolongation (Rosenberg et al; HCQ) | 144 | 59 | 85 (30, 140) |  |
| Liver (abnormal LFTs; Chen et al; HCQ) | 267 | 200 | 67 (-279, 412) |  |
| Liver (increased ALT; Tang et al; HCQ) | 14 | 13 | 1 (-4, 4) |  |
| Serious adverse event (Tang et al; HCQ) | 29 | 0 | 29 (-11, 68) |  |

HCQ=hydroxychloroquine; CQ=chloroquine; RD=Risk difference; CI= confidence interval; LFT=liver function test; ^Risk ratio

### 4.1 Benefits

#### 4.1.1 Death: all-cause mortality

A reduction in the risk of death from COVID-19 was considered as a key benefit of treatment with chloroquine or hydroxychloroquine compared to standard of care, placebo or other treatments. Data was available from two studies for which mortality risk was either the primary endpoint or included in the primary composite endpoint [19,20], and one additional study in which mortality risk was a secondary objective [21].

The primary outcome in a retrospective cohort study of 1438 patients hospitalized in New York was in-hospital mortality. The Cox proportional hazard ratio (HR) for in-hospital death between patients who received hydroxychloroquine alone vs. patients who received neither hydroxychloroquine or azithromycin was 1.08 (95% CI 0.63-1.85), whilst the HR for hydroxychloroquine and azithromycin vs neither drug was 1.35 (95% CI 0.76-2.40) [19]. A further observational study compared the incidence of intubation or death (composite outcome) in a time to event analysis between patients who received hydroxychloroquine vs. those patients who did not [20]. The adjusted HR for this composite outcome was 1.00 (95% CI 0.76-1.32).

Mortality risk was a secondary outcome in an observational study in which the primary endpoint was survival without transfer to the ICU at day 21; the risk of death amongst the hydroxychloroquine patients was higher (11%) compared to standard of care (9%) [21].

#### 4.1.2 Intensive Care Unit (ICU) admission

A reduction in the risk of ICU admission was identified as a key benefit; results from three studies contained data relating to ICU admission or mechanical ventilation. One observational study examined the use of hydroxychloroquine at a dose of 600 mg/day within 48 hours of admission to hospital (treatment group) versus standard of care without hydroxychloroquine (control group) amongst patients with SARS-CoV-2 pneumonia. The primary outcome was survival without transfer to the ICU at day 21 (HR 0.8 (95% CI 0.4-1.5)). Secondary outcomes were overall survival, survival without acute respiratory distress syndrome (ARDS), weaning from oxygen, and discharge from hospital to home or rehabilitation (all at day 21). No statistically significant differences were observed between the two groups for any of the primary or secondary outcomes [21]. Results from another study showed the risk of ICU admission and mechanical ventilation were both higher amongst patients taking hydroxychloroquine (with and without azithromycin) compared to control [19]. A further study indicated that the risk of intubation was higher among patients taking hydroxychloroquine compared to standard of care [20].

#### 4.1.3 Non-invasive ventilation/High flow oxygen

Another key benefit identified was reduction in the risk of non-invasive ventilation/high flow oxygen. No data comparing hydroxychloroquine or chloroquine to standard of care, placebo or other treatments were identified.

#### 4.1.4 Secondary clinical outcomes

Secondary clinical outcomes were also considered key benefits as they refer to anticipated clinical endpoints as a result of chloroquine or hydroxychloroquine treatment, which reflect potential reductions in disease progression, such as development of pneumonia or ARDS. The only data available relating to secondary clinical outcomes was for survival without ARDS, which was a secondary endpoint in the study by Mahevas et al (primary endpoint was survival without transfer to the ICU at day 21) [21]. Results suggested that survival without ARDS was similar between both groups (30% hydroxychloroquine group, 26% standard of care) [21].

#### 4.1.5 Oxygen

A further key benefit identified was a reduction in the number of patients requiring supplemental oxygen. Data for this key benefit was only identified in one study. The study by Mahevas et al provided results for oxygen weaning, between the hydroxychloroquine group and standard of care and the findings were similar for both groups (79% and 74%) [21].

#### 4.1.6 Viral load parameters

Viral load parameters were considered a surrogate endpoint in this benefit risk assessment context, with outcomes such as virological clearance reflecting the benefit of recovery from COVID-19. Five studies provided data on this outcome. In the study by Gautret et al [22], the authors examined virological clearance (Polymerase Chain Reaction (PCR) of nasopharyngeal samples) at day six post study inclusion as the primary outcome. Of a total sample size of 36 patients, 20 were treated with hydroxychloroquine and 16 received standard of care (control group). A higher proportion of those in the hydroxychloroquine group had negative PCR results at day six (0.70) compared to those in the control group (0.13; Risk Difference (RD)=0.58, 95% CI: 0.17, 0.98).

The study by Chen et al [23] examined virological clearance (negative conversion rate of COVID-19 nucleic acid from respiratory pharyngeal sample) on day seven after randomisation. Fifteen patients were treated with hydroxychloroquine and 15 patients received standard of care (comparator group). No significant difference in proportion of viral clearance was observed between the hydroxychloroquine group (0.87) and the comparator group (0.93; RD=-0.07, 95% CI: −0.75, 0.61). Median time to clearance was comparable between the two groups (hydroxychloroquine: 4 days (range 1-9 days), comparator: 2 days (range 1-4 days); p>0.05).

Huang et al [24] examined virological clearance (by Reverse Transcriptase-PCR) on day 14 post randomisation. Ten patients were treated with chloroquine and 12 patients were treated with lopinavir/ritonavir (comparator group), which is a recommended treatment for COVID-19 in China [24]. No significant difference in proportion of viral clearance was observed between the chloroquine group (1.00) and the comparator group (0.92; RD=0.08, 95% CI: - 0.74, 0.91).

Tang et al assessed the efficacy and safety of hydroxychloroquine plus standard of care compared with standard of care alone amongst patients with COVID-19 (majority had mild to moderate disease severity) in a randomised controlled trial [25].The primary outcome was the proportion of patients with negative conversion by day 28. Administration of hydroxychloroquine did not result in a significantly higher proportion of negative conversion compared to standard of care (difference between groups was 4.1% (95% CI –10.3% - 18.5%). A further study by Hraiech et al showed that viral clearance (negative SARS-CoV-2 PCR at day 6 from treatment) was lower amongst patients receiving hydroxychloroquine and azithromycin (18%) compared to lopinavir/ritonavir (38%) and standard of care (20%) [26].

### 4.2 Risks

#### 4.2.1 Cardiac

One study compared the incidence of cardiac arrest and abnormal ECG findings (defined as arrhythmia or prolonged QT fraction) as a secondary objective in a retrospective cohort study of patients [19]. The odds ratio (OR) for cardiac arrest amongst patients who received hydroxychloroquine alone vs patients who received neither hydroxychloroquine or azithromycin was 1.91 (95% CI 0.96-3.81) [19], whilst the OR for hydroxychloroquine plus azithromycin vs neither drug was statistically significant at 2.13 (95% CI 1.12-4.05). The odds ratio for abnormal ECG findings for patients who received hydroxychloroquine alone vs patients who received neither hydroxychloroquine or azithromycin was 1.50 (95% CI 0.88-2.58), whilst the OR for patients who received hydroxychloroquine plus azithromycin vs neither drug was 1.55 (95% CI 0.89-2.67).

In EudraVigilance, there were 13 reports of QT prolongation, one report of ventricular arrythmia, one report of Atrioventricular (AV) block, one fatal cardiac arrest and one non-fatal cardiac arrest in patients using hydroxychloroquine for COVID-19 infection. In addition, there were six reports of QT prolongation, one report of tachyarrhythmia, and one report of ventricular tachycardia in patients using chloroquine for COVID-19 infection.

#### 4.2.2 Immune reactions

No comparative data were identified on immune reactions among patients using hydroxychloroquine or chloroquine for COVID-19 treatment.

#### 4.2.3 Ocular

No comparative data on ocular events among patients using hydroxychloroquine or chloroquine for COVID-19 treatment were identified.

#### 4.2.4 Skin

No comparative data were identified on serious skin reactions among patients using hydroxychloroquine or chloroquine for COVID-19 treatment.

#### 4.2.5 Liver

Two studies reported data on liver function outcomes. In the study by Chen J et al [23], abnormal liver function was reported for four patients in the hydroxychloroquine group (risk=0.27) and three patients in the comparator group (risk=0.20). There was no significant difference in risk between the two groups (RD=0.07, 95% CI: −0.28, 0.41). One report of increased alanine aminotransferase was reported amongst both groups (standard of care plus hydroxychloroquine vs. standard of care alone) in the study by Tang et al [25].

In EudraVigilance, there were two reports of liver injury and one report of hepatocellular injury in patients using hydroxychloroquine for coronavirus infection. In addition, there was one report of hepatocellular injury in a patient using chloroquine for COVID-19 infection.

#### 4.2.6 Severe hypoglycaemia

No comparative data were identified on severe hypoglycaemia among patients using hydroxychloroquine or chloroquine for COVID-19 treatment.

#### 4.2.7 Blood

No comparative data were identified for adverse haematological events. In EudraVigilance, there were two reports of neutropenia in patients using hydroxychloroquine for COVID-19 infection.

#### 4.2.8 Drug interactions

No comparative data were identified on drug interactions with chloroquine or hydroxychloroquine. In EudraVigilance, one potentiating drug interaction was reported in a patient using chloroquine for COVID-19 infection; the patient experienced QT prolongation and was also taking lithium carbonate and quetiapine fumarate.

## 5 Discussion

Several potential key benefits and risks were identified with use of hydroxychloroquine or chloroquine in COVID-19 treatment and included in the value tree. A limited number of studies were identified that compared benefits and risks between those using hydroxychloroquine or chloroquine and those receiving standard of care, placebo or other treatments, hence data were identified for only some of these key benefits and risks at this current time.

Amongst the three studies which reported on mortality risk, none showed a reduced risk amongst patients receiving chloroquine or hydroxychloroquine. With respect to ICU admission, no statistically significant difference in the risk of ICU admission was demonstrated [21], whereas another study showed increased risk of ICU admission and mechanical ventilation amongst patients taking hydroxychloroquine compared to control[19]. Following data lock for this assessment, a statement was released by the chief investigators of the RECOVERY trial on hydroxychloroquine use in hospitalised patients [27]. This is a large randomised clinical trial which has enrolled over 11,000 patients across the UK so far, 1542 of which were randomised to receive hydroxychloroquine [27]. A review of the unblinded data for this arm of the trial revealed no significant difference in the primary endpoint of 28 day mortality between hydroxychloroquine (25.7% of 1542 patients) and usual care (23.5% of 3132 patients; HR=1.11, 95% CI: 0.98, 1.26) [27]. While this data has yet to be published, the initial findings from this trial are concordant with the results of other studies examined in this benefit-risk assessment.

There was a lack of data relating to indicators of disease severity such as the development of pneumonia or ARDS; one study suggested that the proportion of patients who survived without ARDS, and who were weaned off oxygen, were similar between the hydroxychloroquine and standard of care groups [21]. Whilst five studies reported outcomes relating to virological clearance, only one study revealed a significant risk difference between hydroxychloroquine and the comparator group (standard of care), though given the small sample size and biases in the study design this should be interpreted with caution. All five studies examined virological clearance at different time points with different comparators, meaning results from these studies could not be pooled.

In addition, several potential key risks were also identified. Cardiac toxicity is of particular importance for COVID-19 patients. Both the disease itself and anticipated treatment strategies with chloroquine or hydroxychloroquine potentially pose significant risk of cardiac arrhythmias. [28]. Short term use, as expected for the treatment of COVID-19, is likely to pose a lower risk of cardiac toxicity, nevertheless the risk cannot be overlooked as patients are expected to be on higher doses, possibly concomitantly taking other QT prolonging agents, in addition to having a potentially elevated risk due to the disease itself. One study reported a statistically significant OR for the risk of cardiac arrest amongst patients receiving hydroxychloroquine plus azithromycin vs neither drug (OR 2.13 (95% CI 1.12-4.05)) [19], whilst the OR for cardiac arrest amongst patients who received hydroxychloroquine alone vs patients who received neither hydroxychloroquine or azithromycin was non-significant at 1.91 (95% CI 0.96-3.81). Whilst this may be due to the synergistic effect of this drug combination on QT prolongation, Rosenberg et al acknowledged that not all potential confounders were available for modelling purposes, and that unmeasured residual confounding could not be excluded [19].

One of the most serious toxic effects of hydroxychloroquine and chloroquine are ocular side effects, notably retinopathy [29–33], although the risk for hydroxychloroquine is considered to be lower [34]. Both duration of use and weight-based dosing (dose per kg) are important parameters for the risk of developing retinopathy [33]. The risk of retinal damage over a short time period may be negligible even with high doses [32]. However, given the seriousness of the outcome we have identified this as a key risk in the benefit-risk framework, though no comparative data was identified. In addition, whilst the most robust evidence for safety concerns associated with chloroquine and hydroxychloroquine is with longer term use, although rare, there have been case reports after short-term use, with respect to adverse hepatic and haematological effects, and hypoglycaemia [35–37]. No comparative data for severe hypoglycaemia and haematological events were identified, though comparative data from two studies were available for hepatic outcomes. For the risk of abnormal liver function tests (LFTs), two studies reported the incidence of abnormal LFTs in both treatment groups. Both studies had small sample sizes and no significant risk difference between hydroxychloroquine and the comparator group (standard of care) was observed.

Whilst there is a paucity of comparative data from the literature search at this stage, we identified a number of reports in Eudravigilance. The following data was obtained from EudraVigilance for patients taking chloroquine or hydroxychloroquine: QT prolongation (n=19), ventricular arrythmia (n=1), atrioventricular (AV) block (n=1), non-fatal cardiac arrest (n=1), fatal cardiac arrest (n=1), tachyarrhythmia (n=1), ventricular tachycardia (n=1), liver injury (n=2), hepatocellular injury (n=2), neutropenia (n=2), drug interaction (n=1). These data can be used in future benefit-risk assessments once an appropriate treatment comparator has been identified, which should be an established treatment for COVID-19.

Given the limited availability of data at this stage on benefits and risks with hydroxychloroquine or chloroquine compared to standard of care, placebo or other treatments, we chose not to undertake a fully quantitative assessment of the benefit-risk balance at this time using wNCB. Instead, all available comparative data for key benefits and risks are presented in a summary table. Currently, there is insufficient evidence to suggest a favourable benefit-risk profile for chloroquine or hydroxychloroquine compared to standard of care. Most of the available data for the clinical endpoints identified included information relating to risk of death and ICU admission, for which currently available results do not show an improvement with hydroxychloroquine or chloroquine. Further data from clinical trials and observational studies are required in order to determine whether the benefit-risk profile of hydroxychloroquine or chloroquine in treating COVID-19 is favourable.

### 5.1 Strengths and Limitations

Sample sizes for each outcome were limited to those available in the original studies and may not have adequate power to detect differences in risk between groups, especially where the outcomes examined were not the primary outcome of interest. The benefit-risk assessment is limited by the availability of data in the published literature. However, this assessment can be subsequently updated once further data from clinical trials are available. In addition, given the public health urgency of the COVID-19 pandemic, it is important to provide a systematic assessment of the benefits and risks of hydroxychloroquine and chloroquine treatments with evidence available to date and create a framework which can be used to rapidly update the assessment when further data are available.

Data quality is not reflected in this benefit-risk assessment, though all data included were extracted from peer-reviewed manuscripts. Of note, a statement was issued by the International Society of Antimicrobial Chemotherapy [38] regarding the Gautret et al paper published in the international journal of antimicrobial agents [22]. The paper was not considered to meet the society’s expected standards and though it was peer-reviewed, the editor-in-chief was not involved in this process [38]. The paper by Rosenberg et al acknowledged that adverse events may have occurred at any time during hospitalisation regardless of when hydroxychloroquine was initiated, though hydroxychloroquine initiation usually occurred within one day of hospitalisation. A paper published in the Lancet was identified through the original literature search but was subsequently retracted and so data have not been presented [39]. This highlights the dynamic nature of this benefit-risk assessment.

Confirmation of causality was not a requirement for inclusion of data in the BRAT assessment. Patients may have been on other concomitant medications or had other medical conditions at the time of hydroxychloroquine or chloroquine treatment. Where possible, we have specified the use of hydroxychloroquine or chloroquine with or without use of concomitant medications, for clarity. Finally, we considered hospitalisation of patients to reflect severe COVID-19, but we acknowledge that severity of disease may vary regardless of hospitalisation.

### 5.2 Conclusions

Overall, based on the available data there does not appear to be a favourable benefit-risk profile for chloroquine or hydroxychloroquine compared to standard of care in treatment of severe COVID-19. As further data from clinical trials and real world use on these benefits and risks becomes available, this can be incorporated into the framework for an ongoing benefit-risk assessment.

## Data Availability

Data used in this analysis are available from the references supplied.

